# Associations between cord serum antibodies against phosphorylcholine and bacterial infections in neonates: a prospective cohort study in singletons and twins

**DOI:** 10.1101/2024.02.14.24302847

**Authors:** Ruoqing Chen, Yeqi Zheng, Weiri Tan, Feng Wu, Hui Liang, Xi Chen, Youmei Chen, Xian Liu, Fang Fang, Quanfu Zhang, Rui Zhang, Xu Chen

## Abstract

**Background:** Antibodies against phosphorylcholine (anti-PC) are reported to protect against infection. However, the association between cord serum anti-PC and bacterial infection in neonates is yet to be investigated. This study aimed to investigate these associations among both singletons and twins.

**Methods:** A total of 1007 neonates (329 singletons and 678 twins) within the hospital-based Shenzhen Baoan Birth & Twin cohort were included in this study. Levels of IgM anti-PC, IgG anti-PC, as well as IgM, IgG, and IgA in cord serum were measured by enzyme-linked immunosorbent assay. Diagnoses of bacterial infections were identified within 0-27 days after birth. Multivariable logistic regression with propensity score adjustment was performed to assess the associations between levels of antibodies and neonatal bacterial infections.

**Results:** The mean (standard deviation) levels of IgM and IgG anti-PC were 46.68 (14.15) ng/ml and 73.68 (30.44) ng/ml, respectively. Neonatal bacterial infections were diagnosed in 24 singletons (7.29%) and 48 twins (7.08%). A higher level of IgM anti-PC was associated with a lower risk of neonatal bacterial infections in the analyses of singletons (Odds ratio [OR]: 0.64, 95% confidence interval [CI]: 0.41-0.99) or discordant twin pairs (concerning bacterial infection) (OR: 0.44, 95% CI: 0.20-0.95). Statistically significant association was also shown for IgG among singletons and the first-born twins, but not for IgG anti-PC, IgM, or IgA.

**Conclusion:** A higher cord serum level of IgM anti-PC is associated with a lower risk of bacterial infections in neonates.

**Key point:** A higher level of IgM anti-PC in cord serum is associated with a lower risk of bacterial infection in both singleton and twin neonates.

## BACKGROUND

Infection is the primary cause of death among neonates worldwide, especially in developing countries [1]. In China, the standardized neonatal mortality rate was 6.22 per 100,000 live births in 2019 [2]. Additionally, neonatal sepsis and other infections among neonates exhibited the highest standardized years lived with disability in 2019, representing a significant increase compared to 1990 (117.08 versus 15.58 per 100,000 live births) [2].

Phosphorylcholine (PC), a phosphorylated choline derivative, is found on oxidized low-density lipoprotein in atherosclerotic lesions, deceased cells, and several pathogens such as *Streptococcus pneumoniae* [3] and *Staphylococcus aureus* [4]. For microorganisms, PC could be synthesized using choline from the host and serve as a resource of nutrients and an osmoprotectant [5, 6]. Moreover, PC allows bacteria to evade host immune responses and subsequently colonize in the upper respiratory tract [7, 8]. Antibodies against PC (anti-PC) were initially recognized for their anti-infective properties [9] and were later reported to be a protection marker of cardiovascular disease [10], autoimmune diseases [11], as well as fetal growth and premature birth [12]. However, to the best of our knowledge, few studies have assessed the associations between anti-PC and bacterial infections in neonates.

In this prospective cohort study of singleton and twin neonates, we aimed to investigate the associations between levels of anti-PC and other antibodies in cord serum and the risk of neonatal bacterial infections. We hypothesized that anti-PC are protective for neonates, i.e., a higher level of anti-PC in cord serum is associated with a lower risk of bacterial infections.

## METHODS

### Study population

This study was embedded in the ongoing Shenzhen Baoan Birth & Twin (SZBBTwin) cohort, which was initiated in the Baoan Women’s and Children’s Hospital, Shenzhen University, Shenzhen, China, in May 2020. Until March 2023, 343 singletons and 722 twins (361 twin pairs) born through cesarean section had been recruited. Medical information was collected from the hospital information system, and biological samples were collected at the time of birth (including cord blood, amniotic fluid, tissues of placenta and umbilical cord, as well as maternal blood). Neonates without cord blood samples were excluded, resulting in 329 singletons and 678 twins in this study (Figure 1). This study and SZBBTwin cohort were approved by the Ethics Committee of Baoan Women’s and Children’s Hospital (the present study: LLSC2020-03-12; SZBBTwin: LLSC2019-02-18). Informed consent was obtained from the mothers before delivery.

**Figure 1.**
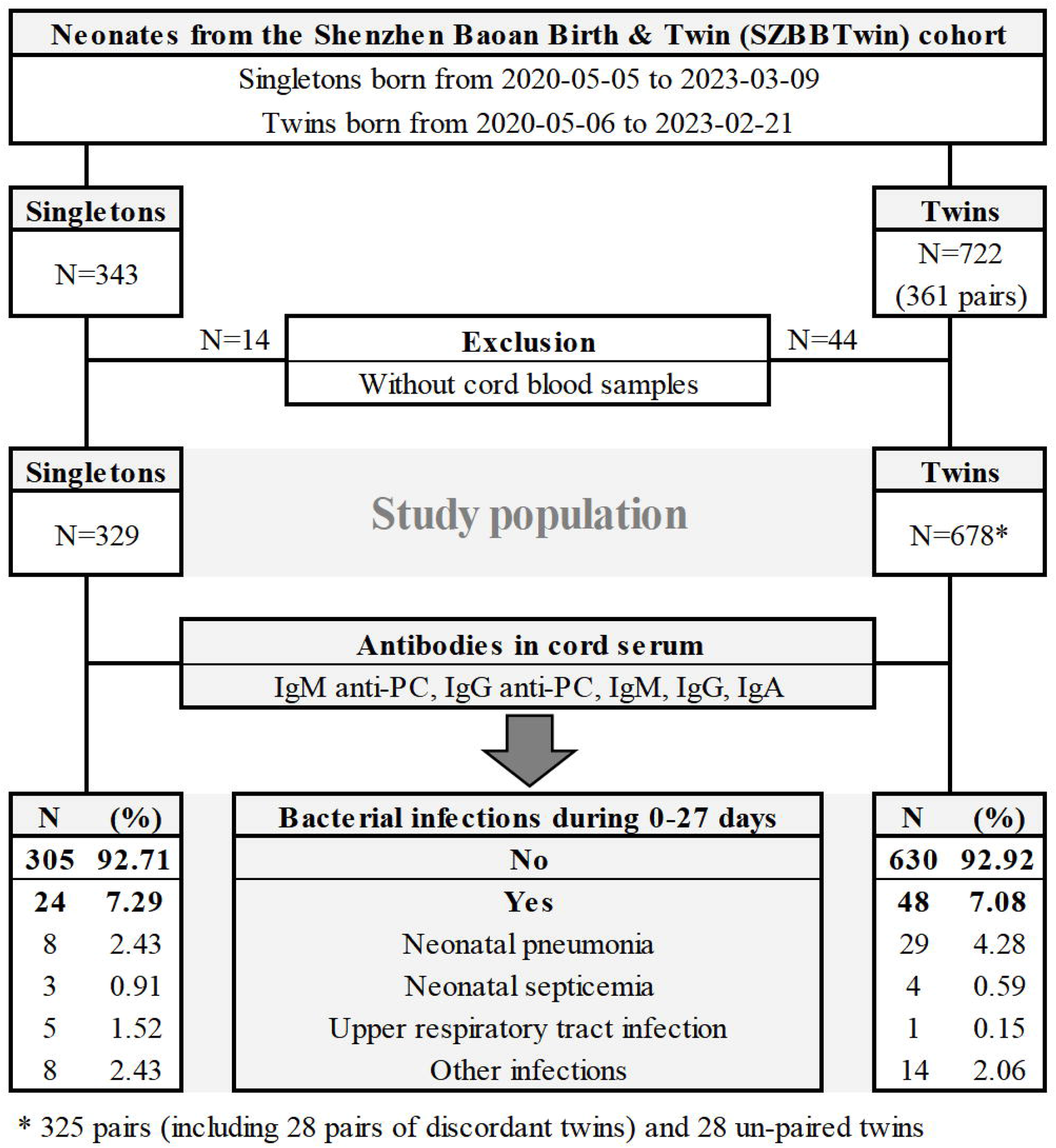
Flow chart of inclusion and exclusion of the study population.

### Measurement of antibodies

The primary exposures were levels of IgM anti-PC and IgG anti-PC in cord serum. To explore the specificity of the associations between anti-PC and neonatal bacterial infections, the total IgM, IgG, and IgA levels were also measured. Cord blood samples were collected during cesarean section, and serum was separated within six hours and stored at -80°C. The five types of antibodies were measured by enzyme-linked immunosorbent assay (ELISA) kits (Jianglaibio Co., Ltd., Shanghai, China) (details in Supplementary Table 1). The cord serum samples underwent first equilibration to room temperature, after which, 50uL of each sample were diluted into 250uL test samples (including 200ul diluent). A double-antibody sandwich ELISA was performed according to the manufacturer’s instructions: 1) 50uL of test samples (penta diluted), calibrators (standard samples), and blank controls (diluent) were dispensed in wells on the pre-coated plates (96 wells); the plates were then sealed and incubated at 37°C for 60 minutes; 2) the contents in wells were aspirated, and 100uL of Horseradish peroxidase conjugated antibody was added in each well; then the plates were sealed and incubated at 37°C for 60 minutes; 3) all contents were aspirated, and 350uL of 1× wash buffer was added and then removed after one minute’s incubation (this washing step was repeated for five times); 4) 100uL

Trimethylbenzene substrate was added in each well, and then incubated at 37°C for 15 minutes; and 5) 50uL stop solution was added in each well, and the optical density (OD, absorbance at 450nm) was measured within 15 minutes (RT-6100, Rayto Life and Analytical Sciences Co.,Ltd., Shenzhen, China). For each plate, the standard curve was generated using linear regression, in which the concentrations and OD values of calibrators (standard samples) were plotted on the X and Y axis, respectively. The concentrations of antibodies in test samples were quantified using the curve equation, and the actual concentrations in cord serum were calculated with dilution factor (test value ×5).

### Neonatal bacterial infection

Neonatal bacterial infection was defined as a diagnosis of bacterial infection within 0-27 days of age. Information on diagnoses was derived from medical records from hospital admissions, outpatient visits, and emergency visits. Additionally, results of laboratory tests, including blood culture, sputum culture, complete blood count, procalcitonin, and C-reactive protein, were used to validate the presence of bacterial infections. Bacterial infection was identified by one of the following criteria: positive sputum culture, positive blood culture, elevated levels of infection indicators in complete blood count (white blood cell count and neutrophil count), procalcitonin>0.05 ng/ml, or C-reactive protein>10 mg/L. For diagnoses during outpatient visits, the presence of purulent secretions observed at physical examination and prescriptions of antibiotics were additionally taken into account.

### Covariates

Maternal and neonatal characteristics were collected from the hospital information system. These characteristics included maternal age at delivery [13], parity [14], educational level [15], mode of conception [16], pre-pregnancy body mass index (BMI) [17], vaginal bleeding during early pregnancy [18], diabetic diseases [19], hypertensive diseases [20], infectious diseases during pregnancy [21], anemia during pregnancy [22], and gestational age at delivery [21]. Characteristics of neonates included sex [23] and birth weight [21].

For twin neonates, information on chorionicity and zygosity was also collected. Chorionicity was determined by ultrasound examinations during the antenatal visit at 11-13 gestational weeks. Opposite-sex twin pairs were considered as dizygotes. For the same-sex twin pairs, DNA was extracted from the cord blood using whole blood genomic DNA purification kits and automatic instrument (MagaBio Plus and NPA-32P, Bioer Technology Co., Ltd., Hangzhou, China). The short tandem repeat genotyping (36A ID System, Microread Genetics Co., Ltd., Beijing, China) was further used to identify the zygosity of same-sex twin pairs.

### Statistical analyses

To address batch effects and ensure that levels of antibodies followed a normal distribution for analyses, levels of all antibodies were adjusted for batch variations by computing residuals based on assay kit batches, followed by rank transformation. Continuous variables were described using mean and standard deviation (SD), and the means of different groups were compared by the Student’s t-test. Categorical variables were described using frequencies and proportions, with group comparisons being performed using the Chi-squared test.

Multivariable logistic regression was used to assess the associations of anti-PC and other antibodies with the risk of neonatal bacterial infections. Model 1 was a crude model. To increase the efficiency of confounding adjustment, we computed a propensity score by modeling a linear regression, with the level of a specific antibody as the dependent variable, and maternal characteristics, including age at delivery, parity, educational level, mode of conception, pre-pregnancy BMI, vaginal bleeding in early pregnancy, diabetic diseases, hypertensive diseases, infectious diseases during pregnancy, anemia during pregnancy, and gestational age at delivery, as well as sex and birth weight of the neonates as the independent variables. In the analyses for singletons, Model 2 adjusted for the propensity score. For analyses of twins, chronicity and zygosity were additionally considered based on Model 2 (Model 3).

To explore the potential impact of birth order on the levels of specific antibodies and neonatal bacterial infection, we performed separate analyses for the first-born twins and second-born twins. To control for unmeasured genetic and environmental factors shared by twins, the analyses were repeated in a sub-sample of twin pairs discordant for neonatal bacterial infection, i.e., discordant twin pairs, using conditional logistic regression (only adjusted for sex and birth weight of the neonates, Model 4).

## RESULTS

In this study, a total of 1007 neonates (329 singletons and 678 twins) were included. Maternal and neonatal characteristics at baseline are shown in Table 1 and Table 2. Neonatal bacterial infections, including pneumonia, septicemia, upper respiratory tract infection, and other infections, were observed in 24 (7.29%) singleton and 48 (7.08%) twin neonates (Figure 1).

**Table 1.**
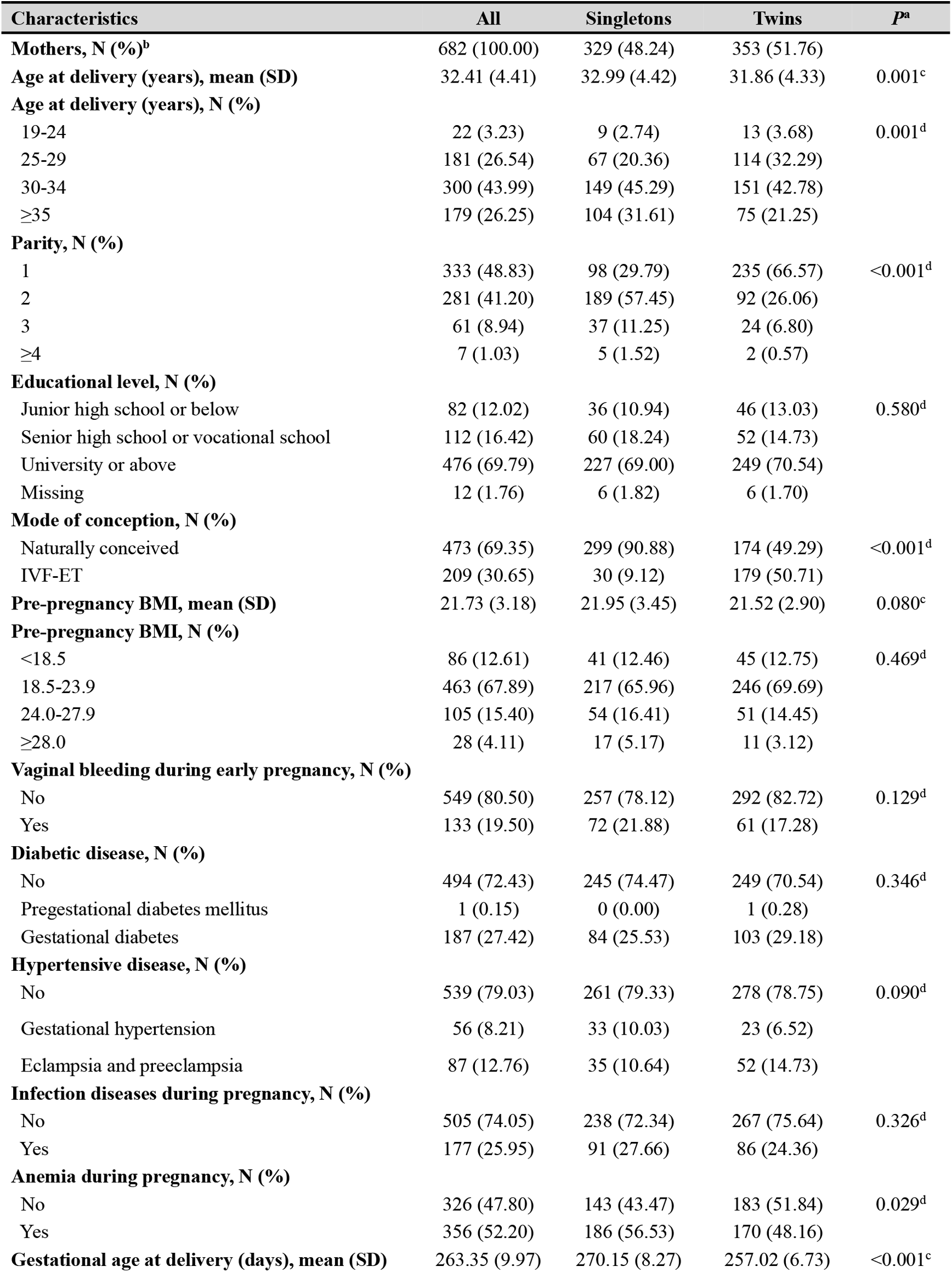
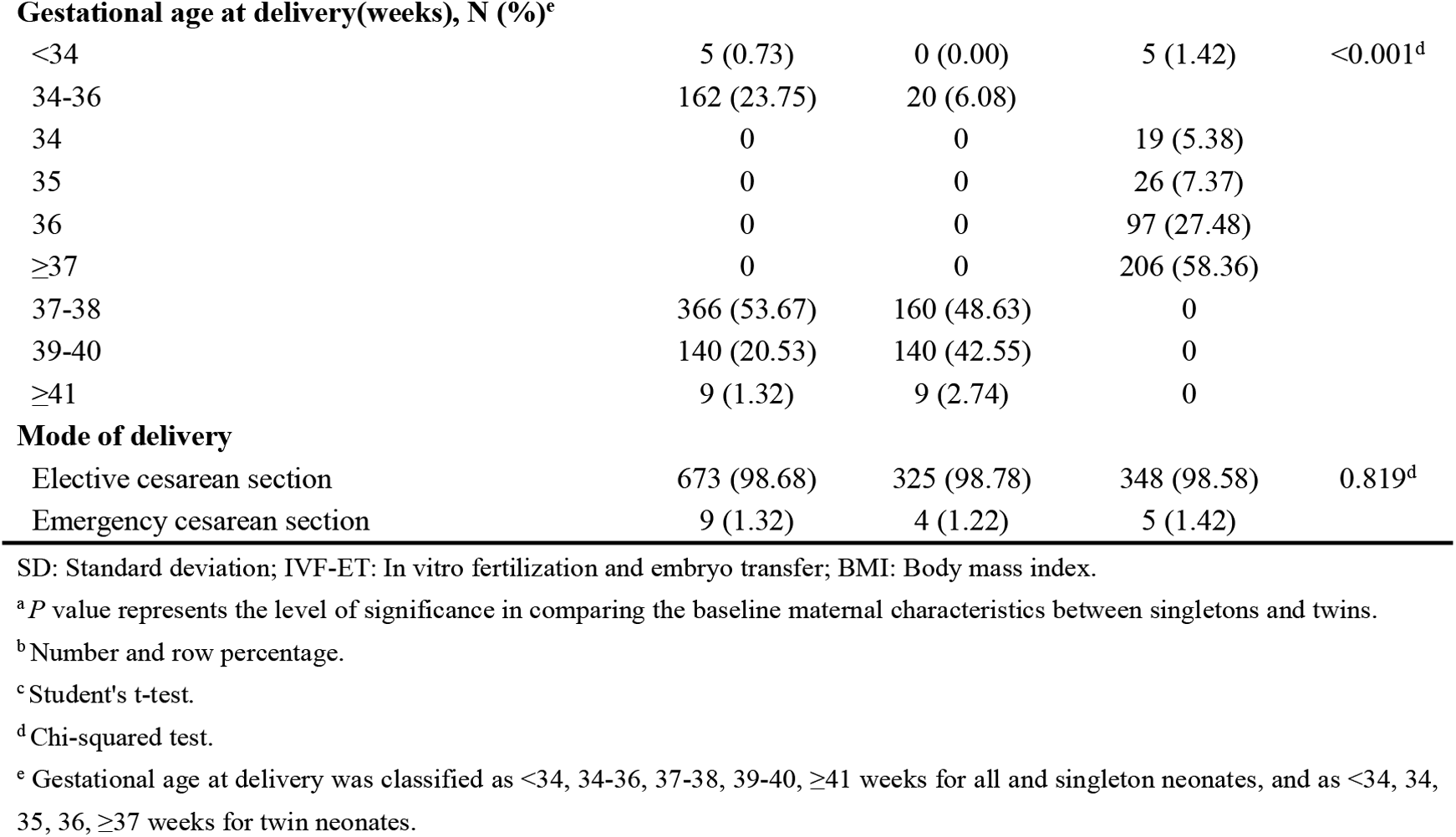
Maternal characteristics at baseline of the study.

| Characteristics | All | Singletons | Twins | *P*<sup>a</sup> |
| --- | --- | --- | --- | --- |
| **Mothers, N (%)<sup>b</sup>** | 682 (100.00) | 329 (48.24) | 353 (51.76) |  |
| **Age at delivery (years), mean (SD)** | 32.41 (4.41) | 32.99 (4.42) | 31.86 (4.33) | 0.001<sup>c</sup> |
| **Age at delivery (years), N (%)** |  |  |  |  |
| 19-24 | 22 (3.23) | 9 (2.74) | 13 (3.68) | 0.001<sup>d</sup> |
| 25-29 | 181 (26.54) | 67 (20.36) | 114 (32.29) |  |
| 30-34 | 300 (43.99) | 149 (45.29) | 151 (42.78) |  |
| ≥35 | 179 (26.25) | 104 (31.61) | 75 (21.25) |  |
| **Parity, N (%)** |  |  |  |  |
| 1 | 333 (48.83) | 98 (29.79) | 235 (66.57) | <0.001<sup>d</sup> |
| 2 | 281 (41.20) | 189 (57.45) | 92 (26.06) |  |
| 3 | 61 (8.94) | 37 (11.25) | 24 (6.80) |  |
| ≥4 | 7 (1.03) | 5 (1.52) | 2 (0.57) |  |
| **Educational level, N (%)** |  |  |  |  |
| Junior high school or below | 82 (12.02) | 36 (10.94) | 46 (13.03) | 0.580<sup>d</sup> |
| Senior high school or vocational school | 112 (16.42) | 60 (18.24) | 52 (14.73) |  |
| University or above | 476 (69.79) | 227 (69.00) | 249 (70.54) |  |
| Missing | 12 (1.76) | 6 (1.82) | 6 (1.70) |  |
| **Mode of conception, N (%)** |  |  |  |  |
| Naturally conceived | 473 (69.35) | 299 (90.88) | 174 (49.29) | <0.001<sup>d</sup> |
| IVF-ET | 209 (30.65) | 30 (9.12) | 179 (50.71) |  |
| **Pre-pregnancy BMI, mean (SD)** | 21.73 (3.18) | 21.95 (3.45) | 21.52 (2.90) | 0.080<sup>c</sup> |
| **Pre-pregnancy BMI, N (%)** |  |  |  |  |
| <18.5 | 86 (12.61) | 41 (12.46) | 45 (12.75) | 0.469<sup>d</sup> |
| 18.5-23.9 | 463 (67.89) | 217 (65.96) | 246 (69.69) |  |
| 24.0-27.9 | 105 (15.40) | 54 (16.41) | 51 (14.45) |  |
| ≥28.0 | 28 (4.11) | 17 (5.17) | 11 (3.12) |  |
| **Vaginal bleeding during early pregnancy, N (%)** |  |  |  |  |
| No | 549 (80.50) | 257 (78.12) | 292 (82.72) | 0.129<sup>d</sup> |
| Yes | 133 (19.50) | 72 (21.88) | 61 (17.28) |  |
| **Diabetic disease, N (%)** |  |  |  |  |
| No | 494 (72.43) | 245 (74.47) | 249 (70.54) | 0.346<sup>d</sup> |
| Pregestational diabetes mellitus | 1 (0.15) | 0 (0.00) | 1 (0.28) |  |
| Gestational diabetes | 187 (27.42) | 84 (25.53) | 103 (29.18) |  |
| **Hypertensive disease, N (%)** |  |  |  |  |
| No | 539 (79.03) | 261 (79.33) | 278 (78.75) | 0.090<sup>d</sup> |
| Gestational hypertension | 56 (8.21) | 33 (10.03) | 23 (6.52) |  |
| Eclampsia and preeclampsia | 87 (12.76) | 35 (10.64) | 52 (14.73) |  |
| **Infection diseases during pregnancy, N (%)** |  |  |  |  |
| No | 505 (74.05) | 238 (72.34) | 267 (75.64) | 0.326<sup>d</sup> |
| Yes | 177 (25.95) | 91 (27.66) | 86 (24.36) |  |
| **Anemia during pregnancy, N (%)** |  |  |  |  |
| No | 326 (47.80) | 143 (43.47) | 183 (51.84) | 0.029<sup>d</sup> |
| Yes | 356 (52.20) | 186 (56.53) | 170 (48.16) |  |
| **Gestational age at delivery (days), mean (SD)** | 263.35 (9.97) | 270.15 (8.27) | 257.02 (6.73) | <0.001<sup>c</sup> |
| **Gestational age at delivery(weeks), N (%)<sup>e</sup>** |  |  |  |  |
| <34 | 5 (0.73) | 0 (0.00) | 5 (1.42) | <0.001<sup>d</sup> |
| 34-36 | 162 (23.75) | 20 (6.08) |  |  |
| 34 | 0 | 0 | 19 (5.38) |  |
| 35 | 0 | 0 | 26 (7.37) |  |
| 36 | 0 | 0 | 97 (27.48) |  |
| ≥37 | 0 | 0 | 206 (58.36) |  |
| 37-38 | 366 (53.67) | 160 (48.63) | 0 |  |
| 39-40 | 140 (20.53) | 140 (42.55) | 0 |  |
| ≥41 | 9 (1.32) | 9 (2.74) | 0 |  |
| **Mode of delivery** |  |  |  |  |
| Elective cesarean section | 673 (98.68) | 325 (98.78) | 348 (98.58) | 0.819<sup>d</sup> |
| Emergency cesarean section | 9 (1.32) | 4 (1.22) | 5 (1.42) |  |

SD: Standard deviation; IVF-ET: In vitro fertilization and embryo transfer; BMI: Body mass index.
<sup>a</sup> *P* value represents the level of significance in comparing the baseline maternal characteristics between singletons and twins.
<sup>b</sup> Number and row percentage.
<sup>c</sup> Student's t-test.
<sup>d</sup> Chi-squared test.
<sup>e</sup> Gestational age at delivery was classified as <34, 34-36, 37-38, 39-40, ≥41 weeks for all and singleton neonates, and as <34, 34, 35, 36, ≥37 weeks for twin neonates.

**Table 2.**
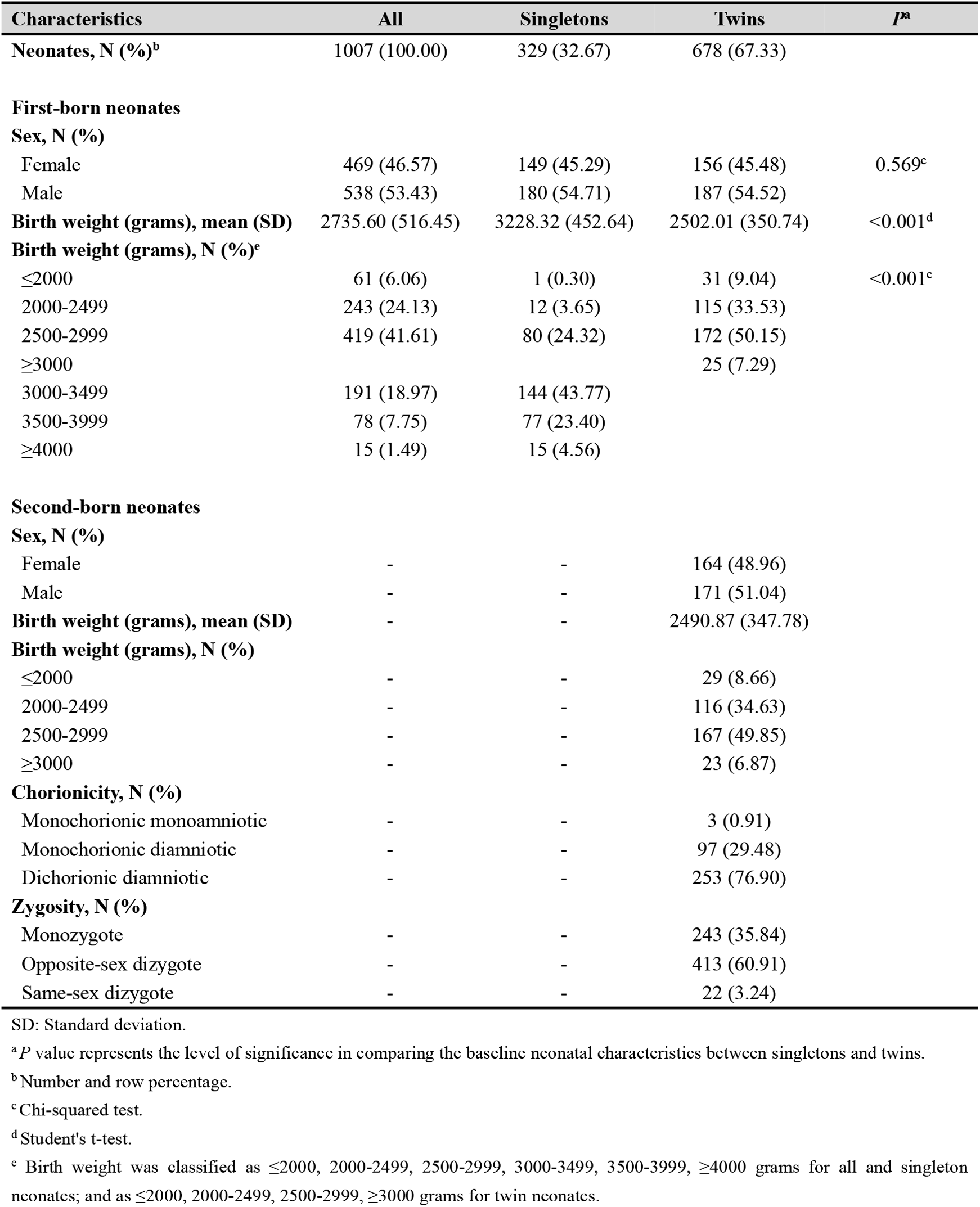
Neonatal characteristics at baseline of the study.

| Characteristics | All | Singletons | Twins | *P*<sup>a</sup> |
| --- | --- | --- | --- | --- |
| **Neonates, N (%)<sup>b</sup>** | 1007 (100.00) | 329 (32.67) | 678 (67.33) |  |
| **First-born neonates** |  |  |  |  |
| **Sex, N (%)** |  |  |  |  |
| Female | 469 (46.57) | 149 (45.29) | 156 (45.48) | 0.569<sup>c</sup> |
| Male | 538 (53.43) | 180 (54.71) | 187 (54.52) |  |
| **Birth weight (grams), mean (SD)** | 2735.60 (516.45) | 3228.32 (452.64) | 2502.01 (350.74) | <0.001<sup>d</sup> |
| **Birth weight (grams), N (%)<sup>e</sup>** |  |  |  |  |
| ≤2000 | 61 (6.06) | 1 (0.30) | 31 (9.04) | <0.001<sup>c</sup> |
| 2000-2499 | 243 (24.13) | 12 (3.65) | 115 (33.53) |  |
| 2500-2999 | 419 (41.61) | 80 (24.32) | 172 (50.15) |  |
| ≥3000 |  |  | 25 (7.29) |  |
| 3000-3499 | 191 (18.97) | 144 (43.77) |  |  |
| 3500-3999 | 78 (7.75) | 77 (23.40) |  |  |
| ≥4000 | 15 (1.49) | 15 (4.56) |  |  |
| **Second-born neonates** |  |  |  |  |
| **Sex, N (%)** |  |  |  |  |
| Female | - | - | 164 (48.96) |  |
| Male | - | - | 171 (51.04) |  |
| **Birth weight (grams), mean (SD)** | - | - | 2490.87 (347.78) |  |
| **Birth weight (grams), N (%)** |  |  |  |  |
| ≤2000 | - | - | 29 (8.66) |  |
| 2000-2499 | - | - | 116 (34.63) |  |
| 2500-2999 | - | - | 167 (49.85) |  |
| ≥3000 | - | - | 23 (6.87) |  |
| **Chorionicity, N (%)** |  |  |  |  |
| Monochorionic monoamniotic | - | - | 3 (0.91) |  |
| Monochorionic diamniotic | - | - | 97 (29.48) |  |
| Dichorionic diamniotic | - | - | 253 (76.90) |  |
| **Zygosity, N (%)** |  |  |  |  |
| Monozygote | - | - | 243 (35.84) |  |
| Opposite-sex dizygote | - | - | 413 (60.91) |  |
| Same-sex dizygote | - | - | 22 (3.24) |  |

SD: Standard deviation.
<sup>a</sup> *P* value represents the level of significance in comparing the baseline neonatal characteristics between singletons and twins.<sup>b</sup> Number and row percentage.<sup>c</sup> Chi-squared test.<sup>d</sup> Student's t-test.<sup>e</sup> Birth weight was classified as ≤2000, 2000-2499, 2500-2999, 3000-3499, 3500-3999, ≥4000 grams for all and singleton neonates; and as ≤2000, 2000-2499, 2500-2999, ≥3000 grams for twin neonates.

Compared with non-infected neonates, the infected neonates were more likely to have mothers who experienced vaginal bleeding during pregnancy and to have been born at a lower gestational age and through emergency cesarean section (Supplementary Table 2). Among twins, neonatal bacterial infections were more likely to occur in twin pregnancies of monochorionic types.

The cord serum levels of IgM anti-PC and IgG anti-PC were on average 46.68±14.15 ng/ml and 73.68±30.44 ng/ml, respectively (Table 3). The cord serum levels of anti-PC were lower in the first-born twins than the second-born twins; the IgM anti-PC levels were 36.99±13.26 ng/ml versus 48.01±11.38 ng/ml, whereas the IgG anti-PC levels were 60.22±23.97 ng/ml versus 88.51±24.13 ng/ml. Similar difference between the first- and second-born twins was also found for total IgM, IgG and IgA. Cord serum levels of anti-PC and other antibodies in relation to maternal and neonatal characteristics in all neonates, singletons, and twins are shown in Supplementary Tables 3 and 4.

**Table 3.** Levels of anti-PC and other antibodies in cord serum among the study population.

| Participants | IgM anti-PC<br>(ng/ml) | IgG anti-PC<br>(ng/ml) | IgM (µg/ml) | IgG (µg/ml) | IgA (µg/ml) |
| --- | --- | --- | --- | --- | --- |
| **All neonates** |  |  |  |  |  |
| Total (N=1007) | 46.68 (14.15) | 73.68 (30.44) | 682.51 (303.15) | 11903.58 (4402.83) | 1831.12 (488.45) |
| Female (N=469) | 46.57 (14.19) | 75.24 (29.67) | 684.12 (298.85) | 11723.43 (4269.60) | 1844.14 (498.66) |
| Male (N=538) | 46.78 (14.13) | 72.32 (31.06) | 681.11 (307.11) | 12060.62 (4513.86) | 1819.78 (479.54) |
| *P*<sup>a</sup> | 0.788 | 0.134 | 0.839 | 0.223 | 0.495 |
| **Singletons** |  |  |  |  |  |
| Total (N=329) | 55.44 (11.03) | 72.61 (35.14) | 697.54 (368.68) | 12236.43 (6096.09) | 1768.84 (584.97) |
| Female (N=149) | 54.31 (10.93) | 73.78 (33.16) | 681.41 (365.25) | 11412.68 (5912.28) | 1787.87 (599.09) |
| Male (N=180) | 56.37 (11.05) | 71.65 (36.76) | 710.89 (371.99) | 12918.31 (6177.58) | 1753.09 (574.23) |
| *P*<sup>a</sup> | 0.091 | 0.522 | 0.643 | 0.021 | 0.309 |
| **Twins** |  |  |  |  |  |
| Total (N=678) | 42.43 (13.53) | 74.20 (27.89) | 675.22 (265.59) | 11742.06 (3274.24) | 1861.35 (431.20) |
| Female (N=320) | 42.96 (14.10) | 75.92 (27.93) | 685.38 (262.93) | 11868.12 (3234.31) | 1870.35 (442.87) |
| Male<br>(N=358) | 41.96 (13.01) | 72.66 (27.80) | 666.13 (267.99) | 11629.38 (3309.98) | 1853.30 (420.96) |
| *P*<sup>a</sup> | 0.400 | 0.111 | 0.387 | 0.291 | 0.551 |
| **First-born twins** |  |  |  |  |  |
| Total (N=343) | 36.99 (13.26) | 60.22 (23.97) | 551.71 (232.52) | 10140.77 (2662.70) | 1656.49 (338.45) |
| Female (N=156) | 36.62 (13.65) | 62.63 (25.54) | 552.39 (226.10) | 10351.45 (2676.86) | 1655.85 (347.83) |
| Male (N=187) | 37.30 (12.95) | 58.21 (22.45) | 551.15 (238.34) | 9965.02 (2645.14) | 1657.02 (331.37) |
| *P*<sup>a</sup> | 0.474 | 0.064 | 0.979 | 0.180 | 0.933 |
| **Second-born twins** |  |  |  |  |  |
| Total (N=335) | 48.01 (11.38) | 88.51 (24.13) | 801.67 (236.47) | 13381.59 (3023.43) | 2071.10 (415.16) |
| Female (N=164) | 49.00 (11.69) | 88.56 (24.01) | 811.89 (231.63) | 13310.81 (3059.86) | 2074.38 (427.65) |
| Male (N=171) | 47.06 (11.04) | 88.46 (24.31) | 791.87 (241.29) | 13449.48 (2995.48) | 2067.95 (404.06) |
| *P*<sup>a</sup> | 0.107 | 0.942 | 0.452 | 0.737 | 0.862 |
| *P*<sup>b</sup> | 0.723 | 0.086 | 0.175 | 0.093 | 0.250 |
| *P*<sup>c</sup> | <0.001 | <0.001 | <0.001 | <0.001 | <0.001 |

Levels were presented as mean (standard deviation).
Anti-PC: Antibodies against phosphorylcholine; IgM: Immunoglobulin M; IgG: Immunoglobulin G; IgA: Immunoglobulin A.
<sup>a</sup> Student's t-test was used to compare the mean levels of antibodies between female and male neonates.
<sup>b</sup> Student's t-test was used to compare the mean levels of antibodies between twins and singletons.
<sup>c</sup> Student's t-test was used to compare the mean levels of antibodies between the first-born and the second-born twins.

After adjustment for propensity score (Table 4), a higher IgM anti-PC level was associated with a lower risk of bacterial infection in the analyses of singletons (odds ratio (OR) 0.64, 95% confidence interval (CI): 0.41-0.99) or discordant twin pairs (OR: 0.44, 95% CI: 0.20-0.95). A higher level of IgG was also associated with a low risk of bacterial infection among singletons and the first-born twins, regardless of adjustment for propensity score. However, such association was not observed among the second-born twins or in the analysis of discordant twin pairs. No association was shown for levels of IgG anti-PC, IgM, or IgA, with the risk of bacterial infections in any groups of neonates.

**Table 4.** Associations of anti-PC and other antibodies with risk of neonatal bacterial infection among the study population.

| Participants | IgM anti-PC (ng/ml) | IgG anti-PC (ng/ml) | IgM (µg/ml) | IgG (µg/ml) | IgA (µg/ml) |
| --- | --- | --- | --- | --- | --- |
| **Singletons (N=329)** |  |  |  |  |  |
| Non-infected, mean (SD) | 55.73 (11.07) | 72.65 (34.80) | 706.48 (369.22) | 12491.51 (6020.97) | 1759.62 (575.56) |
| Infected, mean (SD) | 51.77 (9.99) | 72.10 (40.03) | 583.89 (349.37) | 8994.76 (6243.48) | 1886.03 (697.07) |
| Model 1<sup>a</sup>, OR (95%CI) | 0.67 (0.45-1.01) | 1.00 (0.64-1.58) | 0.82 (0.56-1.20) | 0.61 (0.40-0.92) | 1.01 (0.62-1.65) |
| Model 2<sup>b</sup>, OR (95%CI) | 0.64 (0.41-0.99) | 1.00 (0.62-1.63) | 0.82 (0.55-1.24) | 0.62 (0.39-0.96) | 0.98 (0.59-1.62) |
| **Twins (N=678)** |  |  |  |  |  |
| Non-infected, mean (SD) | 42.66 (13.56) | 73.62 (28.04) | 674.79 (265.88) | 11776.07 (3275.54) | 1856.04 (432.25) |
| Infected, mean (SD) | 39.43 (12.87) | 81.76 (24.81) | 680.88 (264.55) | 11295.71 (3258.17) | 1931.05 (415.26) |
| Model 1<sup>a</sup>, OR (95%CI) | 1.05 (0.79-1.40) | 1.15 (0.90-1.47) | 0.93 (0.69-1.24) | 0.76 (0.57-1.01) | 1.00 (0.78-1.28) |
| Model 3<sup>c</sup>, OR (95%CI) | 1.00 (0.75-1.33) | 1.17 (0.90-1.53) | 0.90 (0.67-1.20) | 0.78 (0.57-1.05) | 0.98 (0.77-1.26) |
| **First-born twins (N=343)** |  |  |  |  |  |
| Non-infected, mean (SD) | 37.33 (13.51) | 59.48 (24.32) | 553.56 (233.38) | 10199.73 (2683.38) | 1650.39 (337.81) |
| Infected, mean (SD) | 32.43 (8.27) | 70.04 (16.19) | 527.17 (224.04) | 9357.09 (2276.07) | 1737.56 (343.78) |
| Model 1<sup>a</sup>, OR (95%CI) | 0.96 (0.62-1.47) | 1.31 (0.83-2.07) | 0.74 (0.45-1.20) | 0.58 (0.35-0.94) | 1.03 (0.62-1.70) |
| Model 3<sup>c</sup>, OR (95%CI) | 1.00 (0.63-1.57) | 1.27 (0.78-2.06) | 0.75 (0.45-1.26) | 0.58 (0.34-0.98) | 1.03 (0.61-1.73) |
| **Second-born twins (N=335)** |  |  |  |  |  |
| Non-infected, mean (SD) | 48.13 (11.27) | 88.12 (23.93) | 799.13 (238.60) | 13392.96 (3033.78) | 2066.97 (417.00) |
| Infected, mean (SD) | 46.43 (12.97) | 93.49 (26.62) | 834.58 (208.60) | 13234.33 (2944.34) | 2124.54 (395.04) |
| Model 1<sup>a</sup>, OR (95%CI) | 1.21 (0.73-2.01) | 1.07 (0.65-1.77) | 1.09 (0.68-1.75) | 0.81 (0.51-1.30) | 0.97 (0.62-1.51) |
| Model 3<sup>c</sup>, OR (95%CI) | 1.03 (0.60-1.78) | 1.28 (0.75-2.18) | 0.99 (0.60-1.64) | 0.82 (0.50-1.34) | 0.87 (0.55-1.39) |
| **Discordant twins (N=56)** |  |  |  |  |  |
| Non-infected, mean (SD) | 47.05 (10.74) | 81.85 (30.70) | 669.64 (290.16) | 12153.50 (3405.26) | 1944.06 (447.85) |
| Infected, mean (SD) | 39.89 (14.60) | 82.63 (27.17) | 716.12 (270.70) | 11136.43 (3096.45) | 1970.14 (394.77) |
| Model 1<sup>a</sup>, OR (95%CI) | 0.57 (0.31-1.03) | 1.04 (0.66-1.63) | 1.13 (0.73-1.77) | 0.78 (0.48-1.26) | 1.07 (0.62-1.84) |
| Model 4<sup>d</sup>, OR (95%CI) | 0.44 (0.20-0.95) | 0.93 (0.55-1.56) | 1.10 (0.67-1.79) | 0.69 (0.39-1.22) | 0.86 (0.45-1.65) |

Anti-PC: Antibody against phosphorylcholine; IgM: Immunoglobulin M; IgG: Immunoglobulin G; IgA: Immunoglobulin A; SD: Standard deviation; OR: Odds ratio; CI: Confidence interval.
<sup>a</sup> Mode 1 is a crude model.
<sup>b</sup> Model 2 adjusted for propensity scores, which were computed from a linear regression model with maternal age at delivery, parity, educational level, mode of conception, prepregnancy body mass index, vaginal bleeding during early pregnancy, diabetic diseases, hypertensive diseases, infection diseases during pregnancy, anemia during pregnancy, gestational age at delivery, and sex and birth weight of the neonates as independent variables and specific antibody as dependent variable.
<sup>c</sup> Model 3 adjusted for propensity scores, which were computed from a linear regression model with variables adjusted in Model 2 plus chronicity and zygosity as independent variables and specific antibody as dependent variable.
<sup>d</sup> Model 4 adjusted for sex and birth weight of the neonates.

## DISCUSSION

In this hospital-based prospective cohort study of singleton and twin neonates, a higher level of IgM anti-PC in cord serum was associated with a lower risk of neonatal bacterial infections in the analyses of singletons or discordant twin pairs. Similar results were found for IgG in singletons and the first-born twins. However, no evidence was observed for associations of IgG anti-PC, IgM, and IgA with neonatal bacterial infections.

We found that a higher cord serum level of IgM anti-PC, not total IgM, was associated with a lower risk of neonatal bacterial infection. PC is a specific epitope on several pathogens, such as *Streptococcus pyogenes* [24], *Haemophilus influenzae* [25], and *Neisseria* [26]. Previous animal experiments suggest that anti-PC may protect the host against pneumococcal infection [27, 28]. Moreover, the presence of anti-PC induced by exposure to PC-containing bacteria underscores its protective role in inhibiting the development of allergic diseases [29]. Therefore, anti-PC may be considered a potential marker for the immune status of neonates.

The association between IgG anti-PC and bacterial infection was not statistically significant in any groups of neonates, but total IgG showed a protective effect on neonatal bacterial infection in singletons. Such pattern is similar to findings from a prospective study of Brazilian twins [30], which showed a higher level of total IgG in cord blood of non-infected neonates than infected neonates. Furthermore, a previous study suggested that cord blood concentrations of serotype III-specific IgG surpassing 7 μg/ml was associated with protection against Group B Streptococci infection in neonates[31], while another study revealed that the majority of neonates with IgG levels below 7 μg/ml would not develop Group B Streptococci infection [32].

Interestingly, we found also an association for IgG in the first-born twins but not in the second-born twins, although no difference was observed in the frequency of bacterial infections between the first- and second-born twins. However, IgG level of the first-born twins was lower than that of second-born twins, suggesting that the first-born twins might be more vulnerable to infection. As for IgA, an Indian study found that IgA concentration in umbilical cord blood might not be a reliable marker for predicting neonatal bacterial infections [33], corroborating the present study.

There are several strengths in our study. First, the cord serum levels of antibodies could reflect directly the initial immune status of neonates [34]. Second, the incorporation of twin design could control for shared genetic and environmental confounding factors between twins, enhancing the internal validity of the study. Third, the use of propensity score adjustment is statistically advantageous in case of small number of outcomes. Nevertheless, some limitations should also be acknowledged. First, we had limited information on the pathogens of neonatal bacterial infections, precluding the possibility of analyzing infections of specific bacteria. Second, collecting cord blood from neonates with small gestational age is challenging, which might have precluded some of the neonates from being enrolled in the SZBBTwin cohort. As a result, the twins in this study might have a larger gestational age than the overall twin population at the hospital who might have higher incidence of neonatal bacterial infection. This, together with the inclusion of only neonates delivered via cesarean section, limits the generalizability of our findings. Third, other biological samples (such as peripheral venous blood) were not collected from neonates, limiting the possibility of studying the dynamic profiles of immunoglobulins in relation to neonatal bacterial infections.

It is worth noting that the study period of the present study included the COVID-19 pandemic when diverse mitigating policies were implemented in China. Coupled with variations in clinical practice in obstetric and neonatal units, this might have introduced additional variation to the results. The implementation of infection control measures, such as enhanced hygiene protocols, visitor restrictions, and changes in hospital workflows, could have influenced the healthcare environment for pregnant women and neonates during this period. These alterations, in turn, might have altered the immune system of neonates, potentially influencing their early life health outcomes, including bacterial infections [35].

In conclusion, our study found that a higher level of IgM anti-PC in cord serum was associated with a lower risk of neonatal bacterial infections among singletons and twins. Additionally, higher IgG levels were associated with a lower risk of such infections among singletons and the first-born twins. However, associations of IgG anti-PC, IgM, and IgA with neonatal bacterial infections were not observed in either singletons or twins. Further studies are required to investigate the mechanisms underlying the associations. The clinical use of IgM anti-PC in preventing neonatal bacterial infections is also worthy with further exploration.

## Data Availability

All data produced in the present study are available upon reasonable request to the authors.

## ACKNOWLEDGEMENTS

The authors would like to thank Dr. Likuan Xiong, Dr. Bin Liu, Dr. Yuanfang Zhu, and Dr. Xu Chen (former president) from Shenzhen Baoan Women’s and Children’s Hospital for the construction of the SZBBTwin cohort.

## Author contributions

Ruoqing Chen and Xu Chen conceptualized the study. Yeqi Zheng, Weiri Tan, and Feng Wu collected and managed the data. Hui Liang and Xi Chen performed the assays. Xian Liu, Youmei Chen, Quanfu Zhang, and Rui Zhang recruited the participants and collected the samples. Yeqi Zheng performed the statistical analyses and wrote the initial draft of the manuscript. Ruoqing Chen, Fang Fang, and Xu Chen revised the manuscript. All authors have read and agreed to the final version of the manuscript.

## Funding sources

This study was funded by the 100 Talents Plan Foundation of Sun Yat-sen University (R. Chen, Grant No.: 58000-12230022), the Guangdong Basic and Applied Basic Research Fund (R. Chen, Grant No.: 2022A1515110417), the Science, Technology and Innovation Council of Shenzhen [X. Chen, Grant No.: JCYJ20190809152801661], and the National Natural Science Foundation of China (X. Chen, Grant No.: 82001652).

## Conflict of interest

The authors declare no conflict of interest.

## Supplementary Data

Supplementary tables are available online.

## Notes

### Competing Interest Statement

The authors have declared no competing interest.

### Author Declarations

Ethics committee of Baoan Women's and Children's Hospital gave ethical approval for this work

